# Characterizing Disparities in the HIV Care Continuum among Transgender and Cisgender Medicare Beneficiaries

**DOI:** 10.1101/2024.03.19.24304525

**Authors:** Jaclyn M.W. Hughto, Hiren Varma, Kim Yee, Gray Babbs, Landon D. Hughes, David R. Pletta, David J. Meyers, Theresa I. Shireman

## Abstract

**Background:** In the US, transgender and gender-diverse (TGD) individuals, particularly trans feminine individuals, experience a disproportionately high burden of HIV relative to their cisgender counterparts. While engagement in the HIV Care Continuum (e.g., HIV care visits, antiretroviral (ART) prescribed, ART adherence) is essential to reduce viral load, HIV transmission, and related morbidity, the extent to which TGD people engage in one or more steps of the HIV Care Continuum at similar levels as cisgender people is understudied on a national level and by gendered subgroups.

**Methods and Findings:** We used Medicare Fee-for-Service claims data from 2009 to 2017 to identify TGD (trans feminine and non-binary (TFN), trans masculine and non-binary (TMN), unclassified gender) and cisgender (male, female) beneficiaries with HIV. Using a retrospective cross-sectional design, we explored within- and between-gender group differences in the predicted probability (PP) of engaging in one or more steps of the HIV Care Continuum. TGD individuals had a higher predicted probability of every HIV Care Continuum outcome compared to cisgender individuals [HIV Care Visits: TGD PP=0.22, 95% Confidence Intervals (CI)=0.22-0.24; cisgender PP=0.21, 95% CI=0.21-0.22); Sexually Transmitted Infection (STI) Screening (TGD PP=0.12, 95% CI=0.11-0.12; cisgender PP=0.09, 95% CI=0.09-0.10); ART Prescribed (TGD PP=0.61, 95% CI=0.59-0.63; cisgender PP=0.52, 95% CI=0.52-0.54); and ART Persistence or adherence (90% persistence: TGD PP=0.27, 95% CI=0.25-0.28; 95% persistence: TGD PP=0.13, 95% CI=0.12-0.14; 90% persistence: cisgender PP=0.23, 95% CI=0.22-0.23; 95% persistence: cisgender PP=0.11, 95% CI=0.11-0.12)]. Notably, TFN individuals had the highest probability of every outcome (HIV Care Visits PP =0.25, 95% CI=0.24-0.27; STI Screening PP =0.22, 95% CI=0.21-0.24; ART Prescribed PP=0.71, 95% CI=0.69-0.74; 90% ART Persistence PP=0.30, 95% CI=0.28-0.32; 95% ART Persistence PP=0.15, 95% CI=0.14-0.16) and TMN people or cisgender females had the lowest probability of every outcome (HIV Care Visits: TMN PP =0.18, 95% CI=0.14-0.22; STI Screening: Cisgender Female PP =0.11, 95% CI=0.11-0.12; ART Receipt: Cisgender Female PP=0.40, 95% CI=0.39-0.42; 90% ART Persistence: TMN PP=0.15, 95% CI=0.11-0.20; 95% ART Persistence: TMN PP=0.07, 95% CI=0.04-0.10). The main limitation of this research is that TGD and cisgender beneficiaries were included based on their observed care, whereas individuals who did not access relevant care through Fee-for-Service Medicare at any point during the study period were not included. Thus, our findings may not be generalizable to all TGD and cisgender individuals with HIV, including those with Medicare Advantage or other types of insurance.

**Conclusions:** Although TGD beneficiaries living with HIV had superior engagement in the HIV Care Continuum than cisgender individuals, findings highlight notable disparities in engagement for TMN individuals and cisgender females, and engagement was still low for all Medicare beneficiaries, independent of gender. Interventions are needed to reduce barriers to HIV care engagement for all Medicare beneficiaries to improve treatment outcomes and reduce HIV-related morbidity and mortality in the US.

## INTRODUCTION

Transgender and gender diverse (TGD) people in the United States (US) are at heightened risk for HIV relative to cisgender (non-transgender) people, with much of the burden experienced by trans feminine people [1–5]. Indeed, a recent analysis of Medicare data found that trans feminine and non-binary (TFN) people had 9 and 4.5 times the probability of having HIV compared to cisgender males and cisgender females or trans masculine and non-binary (TMN) people, respectively [5]. These disparities stem from multilevel sources of stigma that lead to social and economic marginalization, limited access to preventative healthcare, poor mental health, and coping behaviors that can increase the risk of HIV transmission, including injection drug use and sex work [1, 6–9].

The HIV Care Continuum characterizes the HIV care services and outcomes that are essential for reducing HIV-related morbidity, mortality, and forward transmission [10]. These essential health services include linkage and retention in care, as well as uptake and adherence to antiretroviral therapy (ART) to achieve viral suppression [10, 11]. Prior research has documented lower engagement in one or more stages of the HIV care continuum for TGD people, with several studies finding that TFN people have suboptimal ART uptake [12, 13], ART adherence [13–16], and viral suppression [13, 15, 17, 18].

The few studies that have examined differences between TGD people overall relative to cisgender people have found conflicting results. Indeed, one study found that TGD people were less likely than cisgender men and women to be retained in care and achieve viral suppression [18], whereas another study found that TGD people had comparable retention in care as cisgender men, higher ART use than both cisgender men and women, and comparable level of HIV suppression relative to cisgender men and women [19]. Notably, none of these studies comparing TGD people to cisgender people [12–16, 18, 19] explored HIV Care Continuum outcomes among trans masculine and non-binary (TMN) people independent of TFN people, which limits our understanding of important within and between group gender differences. To have a complete understanding of which groups may face barriers to HIV Care Continuum engagement and warrant tailored interventions, national research is needed to explore HIV care engagement among TGD subgroups as well as relative to their cisgender counterparts.

Much of the existing research exploring HIV Care Continuum engagement among TGD people also fails to consider the impact of insurance coverage. This is a critical gap in the literature, given that HIV care quality, satisfaction, and related health outcomes have been shown to differ for members of the general population with and without health insurance [20–23]. Moreover, national survey research finds that 86% of US TGD adults are insured [24] and Medicare insures more than a quarter of people living with HIV/AIDS [5]. Because health insurance is essential to ensuring that people living with HIV can afford the care needed to reduce HIV-related morbidity and mortality [25], understanding TGD and cisgender Medicare beneficiaries’ engagement in the HIV Care Continuum overall and by gender subgroup can identify the subpopulations in greatest need of interventions to optimize HIV care access, use, and delivery.

Building on prior research [5, 26–31], we adapted and applied claims-based methods to achieve the following objectives: [1] identify TGD and cisgender Medicare beneficiaries with HIV and stratify the samples by gender, and [2] explore within- and between-group gender differences in HIV Care Continuum engagement. Findings from this national study can enable a better understanding of HIV Care Continuum engagement among Medicare beneficiaries who are engaged in some level of care, with the goal of identifying the TGD and cisgender subgroups who have the greatest need of tailored interventions to optimize HIV clinical care delivery and improve HIV-related outcomes.

## METHODS

### Study Design/Data Source

We conducted a retrospective cross-sectional analysis to identify US TGD and cisgender Medicare beneficiaries (aged ≥18 years) with HIV and stratify these groups by inferred gender. Fee-for-Service Medicare data were accessed through the Virtual Data Resource Center maintained by the Centers for Medicare & Medicaid Services (CMS). We queried the Medicare Master Beneficiary Summary File and final paid claims for inpatient, physician, and other suppliers, and prescription services from 2008 to 2017. The study was considered exempt by the Brown University Institutional Review Board.

### Identifying TGD Individuals and Stratifying by Gender

To identify TGD individuals, we applied an algorithm that was originally developed with commercial insurance claims data [31] and later adapted for Medicare [5]. These methods and the corresponding codes used to identify the TGD sample are described elsewhere [26, 31]. Briefly, we included any person with a TGD-related diagnosis (e.g., “gender identity disorder”); TGD-conclusive procedures (e.g., “operations for sex transformation, not elsewhere classified”); or a diagnosis of “endocrine disorder not otherwise specified” in conjunction with a transgender-suggestive procedure or gender-affirming hormone prescription.

We subsequently applied a stepwise approach [31] to categorize the inferred gender of the TGD sample. As described in detail elsewhere [5], briefly, we first classified the inferred gender of TGD individuals based on the presence of claims for gender-affirming genital surgeries (e.g., “vaginal construction,” “construction of penis”). Then, within the remaining sample, we categorized the sample by gender if they had certain types of highly specific and highly sensitive reproductive anatomy-specific care and diagnoses (e.g., hysterectomy, pregnancy, prostate cyst, prostate screening). Next, we categorized individuals according to their receipt of gender-affirming hormones or procedures. Finally, using the remaining sample, individuals who had other reproductive anatomy-related diagnoses or procedures (e.g., vulvectomy, testicular hyperfunction) were categorized by inferred gender. These two groups were classified as TMN or TFN. Individuals who had not yet been assigned a gender category or those with conflicting codes at the final step remained unclassified. The unclassified group was comprised of people with a transgender-related diagnosis code and no gender-affirming hormones or procedures or reproductive-anatomy-related care or who had conflicting codes.

### Identifying Cisgender Comparison Group

After identifying the TGD cohort, we selected a random 5% sample from the remaining Medicare beneficiaries, henceforth considered the cisgender sample. The sex of the beneficiaries classified as cisgender was determined from the Master Beneficiary Summary File. Because identification of the TGD cohort relied on engagement in care, we limited the cisgender cohort to beneficiaries who had at least one Part A or B claim and one Part D claim between 2008 and 2017. We also excluded cisgender beneficiaries with missing data on sex and/or date of birth (about 5% of the random sample).

### Identifying Individuals with HIV

We updated an algorithm developed for Medicaid data prior to 2016 and adapted it to identify Medicare beneficiaries with HIV between 2008 and 2017 [29, 30]. First, we used ICD-9 and ICD-10 codes to identify individuals who had an HIV diagnosis code in any claim field (e.g., ICD-9: 043.0 – HIV Disease). Those with a diagnosis code on one or more inpatient or long-term care claims or on two or more claims from an outpatient carrier file were considered to have HIV. Individuals who only had a qualifying diagnosis on one claim from an outpatient carrier file were additionally required to have procedure codes for two or more CD4 count tests, two or more viral load tests, or at least one ART prescription to be considered to have HIV. We identified 1,515 TGD individuals and 11,001 cisgender individuals using HIV diagnosis codes (Electronic Supplement Fig 1 and Fig 2). Next, we identified individuals who were prescribed ART but did not have an HIV diagnosis code documented during the study period. For individuals without an HIV diagnosis, they were first required to have prescriptions for two or more ART ingredients. To ensure that we did not include individuals who were taking antiretroviral medication for HIV prevention (i.e., Pre-Exposure Prophylaxis), we required individuals with no HIV diagnosis and prescriptions for two or more ART ingredients to have diagnostic codes for an HIV-related opportunistic infection (e.g., ICD-10: C46.0-9 - Kaposi Sarcoma), an HIV-related wasting or dementia condition (e.g., ICD-9: 348.3, Encephalopathy, unspecified; ICD-10: R64 - Wasting Syndrome (Cachexia)), or two or more viral load tests or CD4 count tests. The ART pathway identified an additional 303 TGD individuals and 2,220 cisgender individuals (Electronic Supplement Fig 1 and Fig 2). Finally, we identified individuals who were categorized as having HIV within the Chronic Condition Warehouse File and were not captured via the other two pathways (TGD n=253; cisgender n=4,222). Together, these approaches yielded an initial sample of 2,071 TGD and 17,443 cisgender Medicare beneficiaries with HIV. Relatively similar proportions of TGD and cisgender samples were identified within the respective methods, with a modestly higher proportion of TGD people identified using the HIV diagnosis method and a higher proportion of cisgender people identified using Chronic Condition Warehouse indicators (Electronic Supplement Fig 1 and Fig 2).

### Measures

#### Sociodemographics

Age was categorized as 18-24, 25-34, 35-44, 45-54, and 55+ years. These age categories were used because only 10% of the TGD sample with HIV were age 55 years and older at enrollment, and sample sizes for some TGD subgroups (i.e., TMN in the oldest age category) would be too small to report if we split the age categories at 65+. Race and ethnicity were categorized as White (non-Hispanic), Black (non-Hispanic), Hispanic, or Another race and ethnicity (non-Hispanic). Due to small cell sizes that cannot be reported according to our Medicare data use agreement, we combined Asian or Pacific Islander, American Indian or Alaska Native, and other (race or ethnicity) [32]. US Census regions included Northeast, Midwest, South, West, and Unknown.

#### HIV Care Continuum

We explored 5 HIV Care Continuum outcomes: HIV care visit engagement, sexually transmitted infections (STI) screening, ART receipt, 90% ART persistence, and 95% ART persistence. For all measures and for each performance period (i.e., calendar year), we excluded individuals with fewer than 6 months of continuous Fee-for-Service coverage or who died during the performance period. All measures were assessed after the date that an individual was initially identified as having HIV.

To assess <u>HIV Care Visit</u> engagement, we adapted a CMS measure to assess the quality of services provided for the primary management of patients with HIV/AIDS [33] for retrospective measurement with Medicare claims data. Specifically, HIV Care Visit engagement was operationalized as the percentage of adult beneficiaries with HIV who had at least two HIV-related medical visits in a calendar year, with a minimum of 60 days between medical visits.

To assess <u>STI Screening</u> engagement, we adapted a CMS measure to assess the quality of services provided for the primary management of patients with HIV/AIDS [34] for retrospective measurement with Medicare claims data. Specifically, STI testing was operationalized as the percentage of adult beneficiaries with HIV who received chlamydia, gonorrhea, and syphilis screenings at least once per calendar year.

Prescribed ART outcome was operationalized as the percentage of adult beneficiaries with HIV who had at least one day of the 3-drug ART regimen in a calendar year [29, 30].

The <u>90% ART Persistence</u> outcome was operationalized as having at least 90% proportion of days covered (PDC) in a calendar year. We generated a day-by-day array of estimated exposure using the record dates of fills/refills and days supplied for each ART prescription. The PDC corresponds to the proportion of days that a person possessed medication within a defined window of observation and is the Pharmacy Quality Alliance’s preferred method for measuring ART persistence (a proxy for adherence) via claims data [35, 36]. The PDC metric accounts for potential gaps (e.g., hospitalizations) as well as overlapping days for early refills [37]. An individual was considered persistent to ART if their PDC for a 3-drug ART regimen was 90% or greater in any calendar year [38]. Although 95% has historically been considered the gold standard of adherence to ART [39], as of 2019, the Pharmacy Quality Alliance set adherence to ≥90% [36].

The <u>95% ART Persistence</u> outcome was also assessed since guidelines for a lower adherence threshold were set after the last year of the study period [36]. Individuals were considered to have 95% ART persistence if their PDC for a 3-drug ART regimen was 95% or greater in any calendar year.

### Analytic Sample

Using beneficiaries’ date of birth at enrollment, we restricted the sample to individuals who were age 18 years or older at enrollment, who were alive for at least 12 months of a single performance period/year, and who had continuous enrollment in Fee-for-Service coverage for at least six months during one or more performance year/period, resulting in a final analytic sample of 12,779 adult Medicare beneficiaries with HIV, of which 1,678 were TGD and 10,681 were cisgender.

### Data Analysis

All analyses were descriptive. We report the demographic characteristics of the TGD and cisgender samples at enrollment, reporting the raw sample size and frequency overall and by gender subgroup. Since the distribution of sociodemographic characteristics varied by gender subgroup (TFN vs. TMN, cisgender male vs. cisgender female), we calculated the predicted probability of each condition while controlling for age at enrollment. To facilitate within- and between-group comparisons, we obtained the predicted probabilities of each HIV Care Continuum outcome by gender subgroups (TFN, TMN, Unclassified [within the TGD cohort], cisgender males, cisgender females), holding age and gender category at their means. All analyses were conducted using SAS 9.4 (SAS Institute, Cary, NC). Notably, the small sample sizes of some TGD subgroups limited our ability to control for other variables.

## RESULTS

### Demographic Characteristics

Table 1 summarizes the demographic characteristics of the sample. Of the 1,678 TGD beneficiaries with HIV in our sample, 62.7% (n=1,053) were categorized as TFN, 10.1% (n=169) as TMN, and the gender could not be inferred and classified for 27.2% (n=456). Of the 10,681 cisgender individuals with HIV in the comparison group 66.9% (n=7,143) were male, and 33.12% (n= 3,538) were female.

**Table 1.** Demographic characteristics of transgender and gender diverse (TGD) (n=1,678) and cisgender (n=10,681) Medicare beneficiaries with HIV overall and by gender subgroup, 2008-2017.

|  | TGD<br>N=1,678 |  |  |  |  |  | Cisgender<br>N=10,681 |  |  |  | Total<br>N=12,359 |  |  |  |
| --- | --- | --- | --- | --- | --- | --- | --- | --- | --- | --- | --- | --- | --- | --- |
|  | TFN<br>n=1,053 |  | TMN<br>N=169 |  | Unclassified<br>N=456 |  | Male<br>N=7,143 |  | Female<br>N=3,538 |  | TGD<br>N=1,678 |  | Cisgender<br>N=10,681 |  |
|  | N | % | N | % | N | % | N | (%) | N | (%) | N | % | N | % |
| **AGE GROUP** |  |  |  |  |  |  |  |  |  |  |  |  |  |  |
| 18-24 | 76 | 7.2% | 21 | 12.4% | 49 | 10.7% | 115 | 1.6% | 200 | 5.7% | 146 | 8.7% | 315 | 2.9% |
| 25-34 | 242 | 23.0% | 49 | 29.0% | 116 | 25.4% | 588 | 8.2% | 528 | 14.9% | 407 | 24.3% | 1116 | 10.4% |
| 35-44 | 389 | 36.9% | 56 | 33.1% | 145 | 31.8% | 1669 | 23.4% | 718 | 20.3% | 590 | 35.2% | 2387 | 22.3% |
| 45-54 | 244 | 23.2% | 29 | 17.2% | 95 | 20.8% | 2294 | 32.1% | 865 | 24.4% | 368 | 21.9% | 3159 | 29.6% |
| >=55 | 102 | 9.7% | 14 | 8.3% | 51 | 11.2% | 2477 | 34.7% | 1227 | 34.7% | 167 | 10.0% | 3704 | 34.7% |
| **RACE OR ETHNICITY** |  |  |  |  |  |  |  |  |  |  |  |  |  |  |
| White, non-Hispanic | 343 | 32.6% | 76 | 45.0% | 149 | 32.7% | 3309 | 46.3% | 1241 | 35.1% | 568 | 33.8% | 4550 | 42.6% |
| Black, non-Hispanic | 534 | 50.7% | 69 | 40.8% | 228 | 50.0% | 2562 | 35.9% | 1654 | 46.7% | 831 | 49.5% | 4216 | 39.5% |
| Hispanic | 151 | 14.3% | 14 | 8.3% | 55 | 12.1% | 991 | 13.9% | 523 | 14.8% | 220 | 13.1% | 1514 | 14.2% |
| Another race or ethnicity | 25 | 2.4% | 10 | 5.9% | 24 | 5.3% | 281 | 3.9% | 120 | 3.4% | 59 | 3.5% | 401 | 3.8% |
| **CENSUS REGION** |  |  |  |  |  |  |  |  |  |  |  |  |  |  |
| Northeast | 214 | 20.3% | 43 | 25.4% | 99 | 21.7% | 1581 | 22.1% | 946 | 26.7% | 356 | 21.2% | 2527 | 23.7% |
| Midwest | 177 | 16.8% | 25 | 14.8% | 68 | 14.9% | 1044 | 14.6% | 504 | 14.2% | 270 | 16.1% | 1548 | 14.5% |
| South | 431 | 40.9% | 56 | 33.1% | 181 | 39.7% | 3011 | 42.2% | 1637 | 46.3% | 668 | 39.8% | 4648 | 43.5% |
| West | 231 | 21.9% | 45 | 26.6% | 108 | 23.7% | 1455 | 20.4% | 412 | 11.6% | 384 | 22.9% | 1867 | 17.5% |
| Missing | 0 | 0.0% | 0 | 0.0% | 0 | 0.0% | 52 | 0.7% | 39 | 1.1% | 0 | 0.0% | 91 | 0.9% |

**Note.** Demographics were assessed at enrollment. TGD=Transgender and gender diverse. TFN= trans feminine and non-binary; TMN=trans masculine and non-binary; Unclassified=gender category could not be determined. Due to small cell sizes that cannot be reported according to our Medicare data use agreement, we combined Asian/Pacific Islander, American Indian/Alaska Native, and other race or ethnicity under the “Another race or ethnicity” category.

Overall, a higher proportion of TGD beneficiaries with HIV were under age 55 (90.0% TGD; 65.3% cisgender), which can be attributed to the fact that 93.4% of the TGD sample was eligible for Medicare due to disability rather than age, compared to 75.1% of the cisgender sample. Additionally, 62.6% of the TGD sample and 53.6% of the cisgender sample were Black or Hispanic. The largest percentage of beneficiaries for both the TGD (39.8%) and cisgender (43.5%) samples lived in the South at enrollment. The average months of enrollment in Fee-for-Service were also fairly comparable across groups (TGD mean=87.4, Standard Deviation (SD)=32.2; cisgender mean=84.67, SD=35.0).

### HIV Care Continuum

Fig 1 presents results from the age-adjusted models for each HIV Care Continuum outcome for the TGD and cisgender samples and gender subgroups (TMN, TFN, Unclassified, cisgender males, cisgender females). Results from unadjusted models and the exact values from the adjusted model are presented in the Electronic Supplement Tables 1 and 2, respectively.

**Fig 1.**
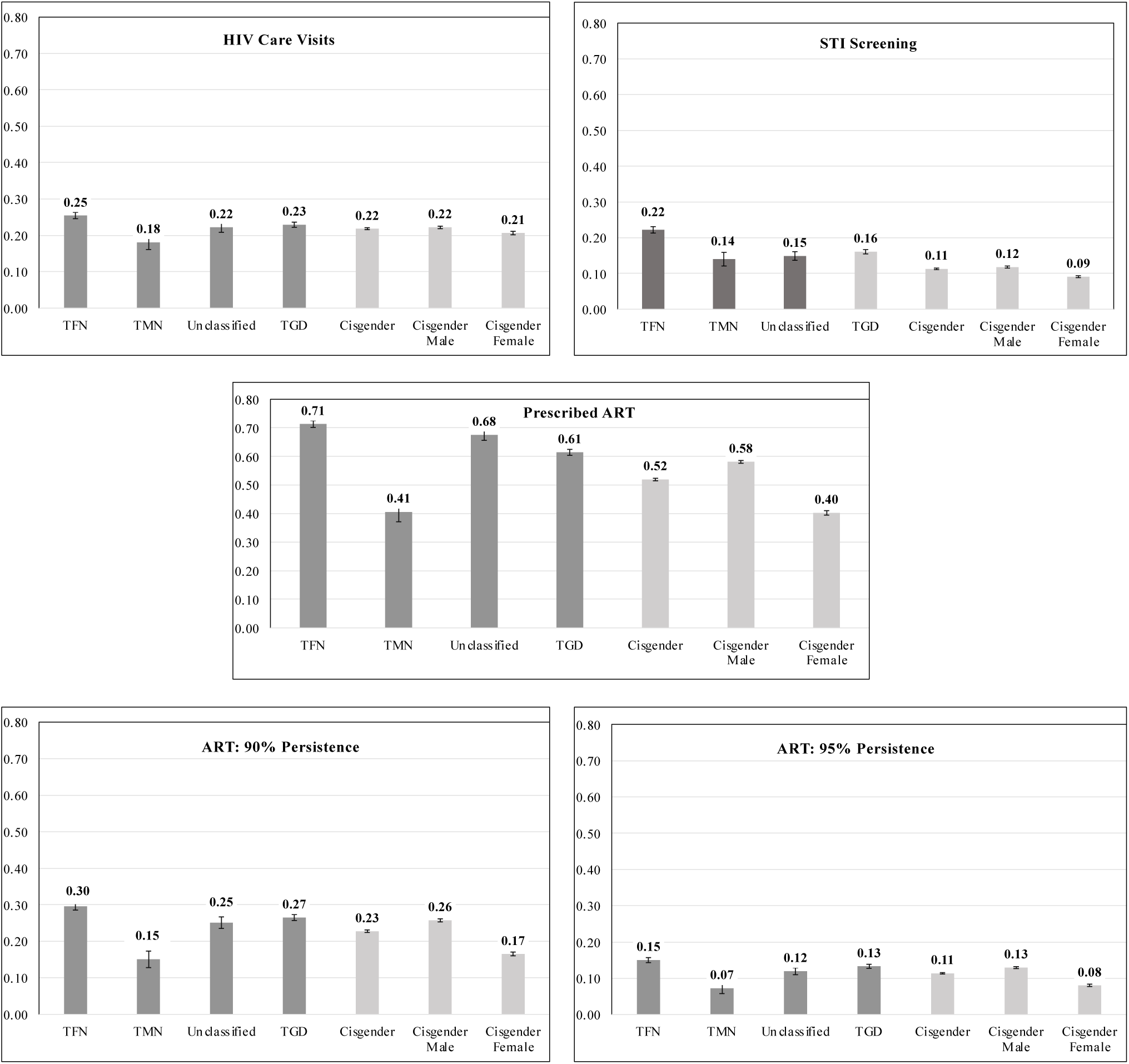
HIV Care Continuum engagement among TGD and cisgender Medicare beneficiaries overall and by gender. The predicted probability of each outcome for TGD (transgender and gender diverse) and cisgender Medicare beneficiaries (2008-2017 by gender: TFN = trans feminine and non-binary; TMN = trans masculine and non-binary; Unclassified = gender category could not be determined. ART = Antiretroviral. STI = Sexually Transmitted Infections.

Once diagnosed with HIV, TGD and cisgender Medicare beneficiaries had a similar predicted probability (PP; herein referred to as “probability) of attending two or more HIV care visits in a given year over the study period (TGD PP=0.22, 95% Confidence Interval (CI)=0.22-0.24; cisgender PP=0.21, 95% CI=0.21-0.22). Moreover, the probability of this outcome was similar across gender subgroups (TFN PP=0.25, 95% CI=0.24-0.27; TMN PP=0.18, 95% CI=0.14-0.22; unclassified PP=0.22, 95% CI=0.20-0.25; cisgender males PP=0.22, 95% CI=0.22-0.23; cisgender females PP=0.21, 95% CI=0.20-0.22).

There were slightly larger differences in the probability of being screened for STIs (i.e., chlamydia, gonorrhea, and syphilis) annually over the study period, with TGD beneficiaries with HIV having a higher probability (PP=0.12, 95% CI=0.11-0.12) compared to cisgender (PP=0.09, 95% CI=0.09-0.10). The probability of annual STI testing was substantially higher among TFN people (PP=0.22, 95% CI=0.21-0.24) relative to all other groups (TMN PP=0.14, 95% CI=0.10-0.18; unclassified PP=0.15, 95% CI=0.13-17; cisgender males PP=0.16, 95% CI=0.15-0.17; cisgender females PP=0.11, 95% CI=0.11-0.12).

For receipt of a 3-drug regimen of ART for at least one day in a calendar year, TGD Medicare beneficiaries with HIV had a higher probability of the outcome (PP=0.61, 95% CI=0.59-0.63) compared to cisgender beneficiaries (PP=0.52, 95% CI=0.52-0.54). This difference is largely driven by the fact that the TFN and the gender unclassified group had a much higher probability of this outcome (TFN PP=0.71, 95% CI=0.69-0.74; unclassified PP=0.68, 95% CI=0.64-0.71).

Ninety percent ART persistence among those prescribed a 3-drug ART regimen were less than half of the probabilities estimated for receipt among TGD (PP=0.27, 95% CI=0.25-0.28) and cisgender people (PP=0.23, 95% CI=0.22-0.23). Further, the probability of 95% persistence was about half the probability for 90% persistence (TGD PP=0.13, 95% CI=0.12-0.14; cisgender PP=0.11, 95% CI=0.11-0.12). For both the 90% and 95% ART persistence outcomes, the probability was meaningfully and significantly highest among TFN people (90% ART Persistence: PP=0.30, 95% CI=0.28-0.32; 95% ART Persistence: PP=0.15, 95% CI=0.14-0.16).

## DISCUSSION

To our knowledge, this is the first national, population-based analysis comparing HIV Care Continuum engagement of TGD and cisgender Medicare beneficiaries with HIV. TGD individuals had a nearly equal or slightly higher predicted probability of every HIV Care Continuum outcome relative to cisgender individuals – a finding that aligns with one prior study showing largely comparable HIV care engagement levels among TGD and cisgender patients initiating care at an HIV clinic [30]. The subgroup differences were more variable, with TFN individuals showing the highest probability of HIV care visit engagement, STI screening, receiving ART, and ART persistence relative to all other groups, which is in opposition to prior studies with individuals of varying or unknown insurance coverage that found lower engagement in one or more stage of the HIV Care Continuum among TFN people relative to cisgender people [12–16]. Further, with the exception of the STI testing, cisgender females and TMN people had a slightly lower probability of engaging in the HIV Care Continuum than TFN people and cisgender males – a finding that extends recent research that showed lower levels of ART adherence among female vs. male Medicare beneficiaries [40]. These findings represent a novel contribution to the field, given that HIV Care Continuum research has not historically included TMN people or reported outcomes for this population separately from TFN people. Despite documented gender differences, it is important to note that HIV care engagement was still low regardless of gender, suggesting potential barriers to HIV care for all Medicare enrollees.

Findings provide insights into the TGD and cisgender groups that could benefit from greater engagement in one or more steps of the HIV Care Continuum. We found comparable probabilities of attending two or more HIV care visits in a calendar year across groups but an elevated probability of being screened for STIs according to HIV quality care guidelines [33, 34] among TGD beneficiaries relative to cisgender beneficiaries. The comparable probability of HIV care visit engagement suggests that regardless of gender, Medicare beneficiaries with HIV are being referred to and/or engaging in HIV care appointments at similar rates, though all groups had low levels of engagement, suggesting barriers to HIV care access and utilization. Conversely, when exploring STI screening, TFN people had the highest probability of meeting the quality care performance measure, followed by TMN individuals. Since TGD people who are on hormones are required to receive lab tests annually [41], it is possible that providers added on STI screening when monitoring hormones. To that end, one recent study found gender-affirming hormone therapy to be associated with a 25% increase in TGD adult patients’ odds of screening for chlamydia and gonorrhea [42]. Future qualitative work with patients and providers is needed to better understand these mechanisms.

We found that TGD beneficiaries were more likely to be prescribed ART in a calendar year than cisgender beneficiaries. Further, we found TGD beneficiaries were more likely to be adherent (or persistent) to ART than cisgender beneficiaries, though 90% and 95% ART persistence were low across all gender groups in alignment with prior research among the general population of Medicare beneficiaries with HIV [40]. Previous work has found mixed results regarding TGD people’s likelihood of ART receipt or any ART use. Some studies found no differences in ART prescription receipt among TFN people compared to other people with HIV [15]. Other studies have found that TGD populations are less likely to receive any ART than cisgender people living with HIV [13]. In contrast, results from studies comparing ART adherence between TGD and cisgender populations have been more consistent. In studies using convenience samples and multi-center cohorts, TGD people have been less likely to be adherent to ART than cisgender people [13–15, 43]. TGD people with HIV have been shown to face barriers to ART adherence, with studies finding that low levels of social support [13], housing instability, and sex work are associated with lower adherence for some TGD populations [44]. Although it is likely that TGD Medicare beneficiaries with HIV in our sample face similar barriers as those with and without insurance who were recruited from HIV clinics and other community sites, it is possible that older and disabled cisgender Medicare beneficiaries face unique barriers to ART adherence [40], thus contributing to suboptimal ART adherence for all people with HIV in our sample.

We utilized claims data from a population insured by Medicare to investigate HIV Care Continuum engagement outcomes, and our findings meaningfully add to knowledge about health insurance and quality care among TGD people living with HIV in the US. To date, many of the studies on US TGD people’s ART use have had limited sample sizes and have not controlled for insurance status [45, 46]. Considering insurance status is critical in light of emerging research showing ART utilization in over 90% of TGD women with HIV living in Canada, a country with universal health coverage [47]. Furthermore, health insurance coverage for gender-affirming hormones may have downstream effects on access to and utilization of ART. In two recent studies, TGD women receiving gender-affirming hormones had higher rates of care retention, ART prescription, and adherence than TGD women not receiving hormones [48, 49]. Further, compared to the general population, TGD people are less likely to have health insurance [50, 51], which may decrease their access to HIV care engagement directly through cost barriers and indirectly through missed linkages with primary and gender-affirming care.

To our knowledge, our study is the first to use Medicare claims data to compare ART prescription and persistence between TGD and cisgender populations. Our study provides fundamental baseline estimates on within- and between-group differences in HIV Care Continuum engagement among a publicly insured adult population. We found evidence of meaningfully higher STI screening and ART persistence within TFN people. Our results support the need for ongoing mixed-methods research to identify the barriers to ART use and persistence for groups with lower levels of utilization (e.g., TMN people and cisgender women) so that tailored interventions can be developed.

### Limitations

The study has limitations. First, since TGD beneficiaries were included in our study based on their observed care, we are unable to validate whether the individuals we identified as TGD were truly so or whether we inaccurately characterized individuals as cisgender based on their absence of qualifying care. However, a similar claims-based algorithm has been found to have high sensitivity and specificity [52], increasing our confidence in our approach. Second, since our data did not capture gender identity, we were forced to infer the gender of our sample and combine individuals who likely hold non-binary and binary gender identities in the same category based on their shared use of certain gender-affirming hormones, procedures, or anatomy-specific care. Non-binary TGD people have been shown to have differential healthcare utilization and risks of various health outcomes than TFN or TMN people [53, 54], and combining these groups may have obscured key differences in healthcare utilization. Third, our algorithm to categorize the gender of TGD people relies on receipt of anatomy-specific or gender-affirming care, so TGD people who did not have claims for this care during the study period or had conflicting claims could not be classified as TFN or TMN. Although the unclassified sample in this study and our prior work [5, 26] have a similar probability of healthcare engagement and chronic conditions (including HIV) as TFN people, it is not possible to discern which gender subgroups comprise this group. Fourth, both TGD and cisgender beneficiaries were included in our study based on their observed care, whereas individuals who did not access relevant care through Fee-for-Service Medicare at any point during the study period were not included. Thus, our sample is unlikely to represent all TGD and cisgender Medicare beneficiaries with HIV. Fifth, our data source did not contain information on important sociodemographic characteristics, such as housing security and income, which may confound the relationship between gender identity and our outcomes. Despite these limitations, our findings provide important new insights into the HIV care received by TGD and cisgender Medicare beneficiaries and serve as a signal for future research and intervention efforts.

## Conclusion

We adapted prior algorithms [5, 26, 29–31] and HIV quality care guidelines [33, 34] to identify a large sample of TGD and cisgender Medicare beneficiaries with HIV and examined within- and between-group differences in HIV Care Continuum engagement among these populations. Our novel methods to identify Medicare beneficiaries with HIV and explore HIV Care Continuum engagement may be helpful for future researchers seeking to study HIV-related outcomes among TGD and cisgender people using claims data. This study also advances HIV health disparities research with TGD and cisgender people [12–16] by documenting within and between group gender differences among TFN, TMN, cisgender male, and cisgender female Medicare beneficiaries with HIV. While our findings regarding TFN people’s engagement differed from some prior studies that did not report insurance status [13, 14, 16] or included uninsured or underinsured people [12, 15], we observed that TGD Medicare beneficiaries had a higher probability of HIV Care Continuum engagement relative to cisgender beneficiaries with HIV, with the highest engagement among TFN people. Although there is a need to increase HIV Care Continuum engagement for all groups to better align with care guidelines, additional mixed-methods research is needed to explore the mechanisms underlying the lower STI screening and ART use documented among TMN people and cisgender females in this sample so that provider and patient-level interventions can be developed to increase engagement and ultimately reduce HIV-related morbidity and mortality among all people living with HIV.

## Data Availability

The data underlying the results presented in the study are available from the Virtual Research Data Center (VRDC) managed by the Centers for Medicare and Medicaid Services. Information on how to access the data and associated fees can be found here: https://www2.ccwdata.org/web/guest/about-vrdc

---

**Supplement Fig 1.**
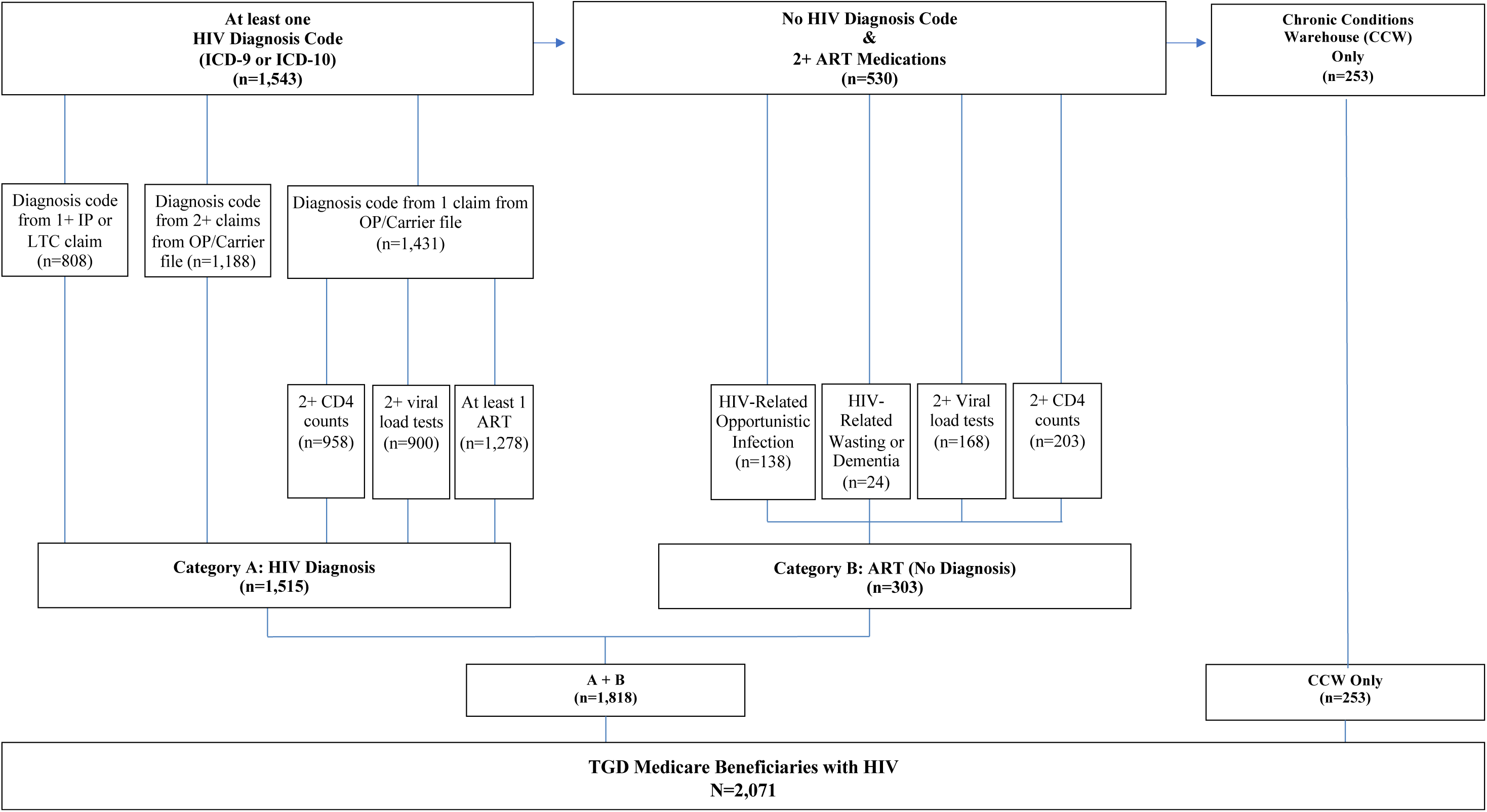
Transgender and Gender Diverse (TGD) Medicare Beneficiaries with HIV, 2008-2017. Flow diagram of TGD sample. The following individuals were excluded: individuals under 18 and those who died or did not have continuous Fee-for-Service coverage for at least 6 months of the performance year, leaving a final analytic sample of N=10,681. IP=inpatient; OP=outpatient; LTC=long term care; ART=antiretroviral medication; ICD=International Classification of Diseases.

**Supplement Fig 2:**
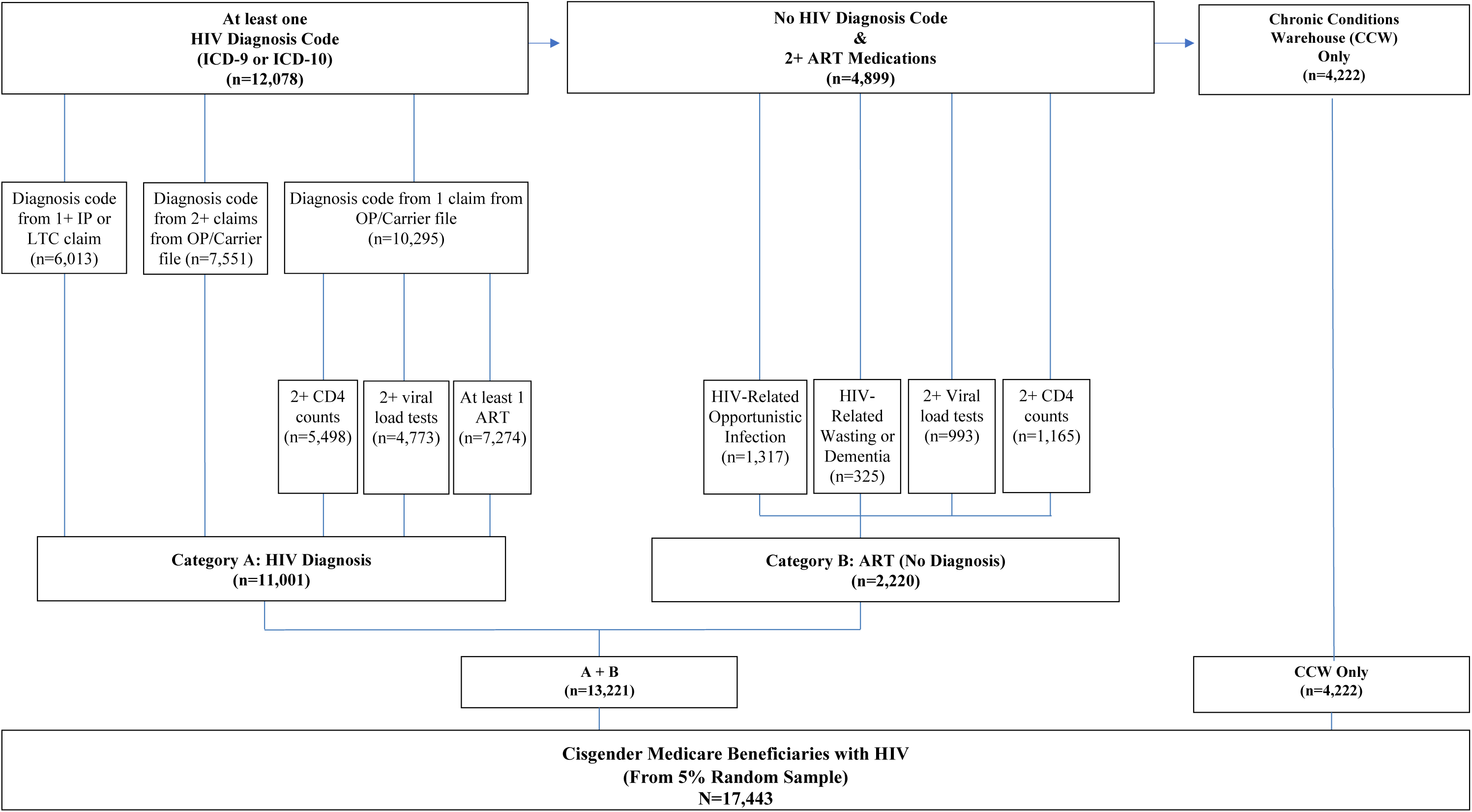
Cisgender Medicare Beneficiaries with HIV, 2008-2017. Flow diagram of TGD sample. The following individuals were excluded: individuals under 18 and those who died or did not have continuous Fee-for-Service coverage for at least 6 months of the performance year, leaving a final analytic sample of N=10,681. IP=inpatient; OP=outpatient; LTC=long term care; ART=antiretroviral medication; ICD=International Classification of Diseases.

**Supplement Table 1.** Unadjusted prevalence of HIV Care Engagement Outcomes among transgender and gender diverse (TGD) (n=1,678) and cisgender (n=10,681) Medicare beneficiaries with HIV, overall and by gender subgroup, 2008-2017.

|  | TGD |  |  |  |  |  | Cisgender |  |  |  | Total |  |  |  |
| --- | --- | --- | --- | --- | --- | --- | --- | --- | --- | --- | --- | --- | --- | --- |
|  | TFN |  | TMN |  | Unclassified |  | Male |  | Female |  | TGD |  | Cisgender |  |
| HIV Care Visits | N=625 | % | N=78 | % | N=232 | % | N=3,719 | % | N=1,747 | % | N=935 | % | N=5,466 | % |
| 2008 | 97 | 19.4% | 12 | 19.4% | 18 | 13.3% | 524 | 16.9% | 273 | 20.2% | 127 | 18.2% | 797 | 17.9% |
| 2009 | 120 | 21.1% | 15 | 18.5% | 37 | 24.0% | 578 | 16.9% | 297 | 18.7% | 172 | 21.4% | 875 | 17.5% |
| 2010 | 146 | 23.1% | 11 | 12.8% | 33 | 17.7% | 668 | 17.7% | 350 | 19.3% | 190 | 21.0% | 1018 | 18.2% |
| 2011 | 153 | 21.8% | 19 | 18.1% | 47 | 21.8% | 782 | 19.1% | 382 | 19.0% | 219 | 21.4% | 1164 | 19.0% |
| 2012 | 176 | 23.1% | 17 | 14.2% | 43 | 17.7% | 848 | 19.2% | 390 | 18.0% | 236 | 21.0% | 1238 | 18.8% |
| 2013 | 213 | 26.7% | 22 | 16.4% | 67 | 24.0% | 1035 | 22.1% | 459 | 19.6% | 302 | 24.9% | 1494 | 21.3% |
| 2014 | 220 | 28.2% | 17 | 13.2% | 64 | 21.8% | 1111 | 23.4% | 472 | 19.8% | 301 | 25.0% | 1583 | 22.2% |
| 2015 | 216 | 28.0% | 25 | 19.5% | 77 | 25.6% | 1223 | 25.0% | 537 | 21.7% | 318 | 26.5% | 1760 | 23.9% |
| 2016 | 242 | 30.7% | 31 | 24.4% | 72 | 22.2% | 1399 | 27.5% | 607 | 23.9% | 345 | 27.8% | 2006 | 26.3% |
| 2017 | 222 | 29.6% | 26 | 20.8% | 87 | 26.0% | 1386 | 26.6% | 581 | 23.1% | 335 | 27.7% | 1967 | 25.4% |
| STI Screening | N=541 | % | N=62 | % | N=168 | % | N=2,028 | % | N=951 | % | N=771 | % | N=2,979 | % |
| 2008 | 63 | 12.6% | 4 | 6.5% | 14 | 10.4% | 192 | 6.2% | 88 | 6.5% | 81 | 11.6% | 280 | 6.3% |
| 2009 | 87 | 15.3% | 8 | 9.9% | 15 | 9.7% | 265 | 7.8% | 128 | 8.1% | 110 | 13.7% | 393 | 7.9% |
| 2010 | 96 | 15.2% | 9 | 10.5% | 16 | 8.6% | 343 | 9.1% | 149 | 8.2% | 121 | 13.4% | 492 | 8.8% |
| 2011 | 134 | 19.1% | 14 | 13.3% | 25 | 11.6% | 400 | 9.8% | 190 | 9.4% | 173 | 16.9% | 590 | 9.7% |
| 2012 | 173 | 22.7% | 16 | 13.3% | 35 | 14.4% | 502 | 11.4% | 177 | 8.2% | 224 | 19.9% | 679 | 10.3% |
| 2013 | 196 | 24.5% | 19 | 14.2% | 41 | 14.7% | 593 | 12.7% | 220 | 9.4% | 256 | 21.1% | 813 | 11.6% |
| 2014 | 187 | 24.0% | 17 | 13.2% | 38 | 12.9% | 617 | 13.0% | 241 | 10.1% | 242 | 20.1% | 858 | 12.0% |
| 2015 | 202 | 26.2% | 25 | 19.5% | 52 | 17.3% | 644 | 13.2% | 259 | 10.5% | 279 | 23.3% | 903 | 12.3% |
| 2016 | 203 | 25.7% | 26 | 20.5% | 71 | 21.9% | 685 | 13.5% | 247 | 9.7% | 300 | 24.2% | 932 | 12.2% |
| 2017 | 194 | 25.9% | 26 | 20.8% | 72 | 21.5% | 751 | 14.4% | 274 | 10.9% | 292 | 24.1% | 1025 | 13.2% |
| Prescribed ART | N=879 | % | N=89 | % | N=365 | % | N=4,838 | % | N=1,759 | % | N=1,333 | % | N=6,597 | % |
| 2008 | 306 | 64.3% | 22 | 37.9% | 88 | 70.4% | 1687 | 54.6% | 514 | 38.0% | 416 | 63.1% | 2201 | 49.5% |
| 2009 | 372 | 68.8% | 27 | 35.5% | 100 | 70.9% | 1898 | 55.6% | 588 | 37.1% | 499 | 65.8% | 2486 | 49.7% |
| 2010 | 439 | 73.3% | 27 | 34.6% | 115 | 69.3% | 2098 | 55.5% | 683 | 37.7% | 581 | 68.9% | 2781 | 49.7% |
| 2011 | 484 | 72.8% | 39 | 40.2% | 139 | 70.2% | 2262 | 55.2% | 739 | 36.7% | 662 | 69.0% | 3001 | 49.1% |
| 2012 | 561 | 77.5% | 44 | 39.3% | 161 | 72.9% | 2596 | 58.8% | 867 | 40.0% | 766 | 72.5% | 3463 | 52.6% |
| 2013 | 592 | 77.2% | 49 | 39.2% | 184 | 71.0% | 2793 | 59.7% | 944 | 40.4% | 825 | 71.7% | 3737 | 53.2% |
| 2014 | 572 | 76.5% | 52 | 43.0% | 200 | 73.5% | 2853 | 60.0% | 991 | 41.6% | 824 | 72.2% | 3844 | 53.9% |
| 2015 | 560 | 75.8% | 54 | 45.0% | 209 | 75.7% | 2945 | 60.3% | 1081 | 43.7% | 823 | 72.5% | 4026 | 54.7% |
| 2016 | 591 | 77.9% | 53 | 44.5% | 224 | 75.9% | 3213 | 63.1% | 1149 | 45.2% | 868 | 74.0% | 4362 | 57.1% |
| 2017 | 576 | 79.8% | 62 | 53.4% | 237 | 78.0% | 3392 | 65.0% | 1205 | 47.8% | 875 | 76.6% | 4597 | 59.4% |
| **ART Persistence: 90%** | **N=576** | **%** | **N=47** | **%** | **N=208** | **%** | **N=3,237** | **%** | **N=1,059** | **%** | **N=831** | **%** | **N=4,296** | **%** |
| 2008 | 94 | 19.7% | 5 | 8.6% | 31 | 24.8% | 619 | 24.0% | 172 | 15.2% | 130 | 19.7% | 791 | 21.3% |
| 2009 | 134 | 24.8% | 10 | 13.2% | 32 | 22.7% | 814 | 28.6% | 225 | 16.7% | 176 | 23.2% | 1,039 | 24.8% |
| 2010 | 163 | 27.2% | 12 | 15.4% | 47 | 28.3% | 917 | 28.9% | 267 | 17.0% | 222 | 26.3% | 1,184 | 25.0% |
| 2011 | 201 | 30.2% | 9 | 9.3% | 39 | 19.7% | 977 | 28.0% | 292 | 16.5% | 249 | 25.9% | 1,269 | 24.2% |
| 2012 | 239 | 33.0% | 14 | 12.5% | 55 | 24.9% | 1,155 | 30.3% | 345 | 17.8% | 308 | 29.1% | 1,500 | 26.1% |
| 2013 | 253 | 33.0% | 22 | 17.6% | 71 | 27.4% | 1,288 | 31.5% | 408 | 19.0% | 346 | 30.1% | 1,696 | 27.2% |
| 2014 | 256 | 34.2% | 20 | 16.5% | 79 | 29.0% | 1,341 | 32.3% | 404 | 18.5% | 355 | 31.1% | 1,745 | 27.6% |
| 2015 | 251 | 34.0% | 25 | 20.8% | 89 | 32.2% | 1,384 | 32.9% | 466 | 20.6% | 365 | 32.2% | 1,850 | 28.6% |
| 2016 | 268 | 35.3% | 21 | 17.6% | 80 | 27.1% | 1,524 | 34.4% | 519 | 22.2% | 369 | 31.5% | 2,043 | 30.2% |
| 2017 | 284 | 39.3% | 28 | 24.1% | 103 | 33.9% | 1,663 | 36.9% | 545 | 23.6% | 415 | 36.3% | 2,208 | 32.4% |
| **ART Persistence: 95%** | **N=429** | **%** | **N=31** | **%** | **N=147** | **%** | **N=2,342** | **%** | **N=726** | **%** | **N=607** | **%** | **N=3,068** | **%** |
| 2008 | 48 | 10.1% | 3 | 5.2% | 14 | 11.2% | 301 | 11.7% | 82 | 7.3% | 65 | 9.9% | 383 | 10.3% |
| 2009 | 74 | 13.7% | 4 | 5.3% | 17 | 12.1% | 383 | 13.5% | 111 | 8.2% | 95 | 12.5% | 494 | 11.8% |
| 2010 | 84 | 14.0% | 6 | 7.7% | 24 | 14.5% | 437 | 13.8% | 140 | 8.9% | 114 | 13.5% | 577 | 12.2% |
| 2011 | 99 | 14.9% | 3 | 3.1% | 19 | 9.6% | 493 | 14.2% | 137 | 7.7% | 121 | 12.6% | 630 | 12.0% |
| 2012 | 124 | 17.1% | 7 | 6.3% | 21 | 9.5% | 599 | 15.7% | 171 | 8.8% | 152 | 14.4% | 770 | 13.4% |
| 2013 | 120 | 15.6% | 8 | 6.4% | 37 | 14.3% | 605 | 14.8% | 183 | 8.5% | 165 | 14.3% | 788 | 12.7% |
| 2014 | 117 | 15.6% | 12 | 9.9% | 42 | 15.4% | 649 | 15.6% | 197 | 9.0% | 171 | 15.0% | 846 | 13.4% |
| 2015 | 132 | 17.9% | 14 | 11.7% | 36 | 13.0% | 658 | 15.6% | 235 | 10.4% | 182 | 16.0% | 893 | 13.8% |
| 2016 | 143 | 18.8% | 14 | 11.8% | 43 | 14.6% | 796 | 18.0% | 270 | 11.6% | 200 | 17.1% | 1,066 | 15.8% |
| 2017 | 142 | 19.7% | 10 | 8.6% | 48 | 15.8% | 818 | 18.1% | 242 | 10.5% | 200 | 17.5% | 1,060 | 15.6% |

**Note.** The percentages reflect the row total for a given year. TGD= Transgender and gender diverse; TFN= trans feminine and non-binary; TMN = trans masculine and non-binary; Unclassified = gender category could not be determined. ART = Antiretroviral. STI= Sexually Transmitted Infections.

**Supplement Table 2.**
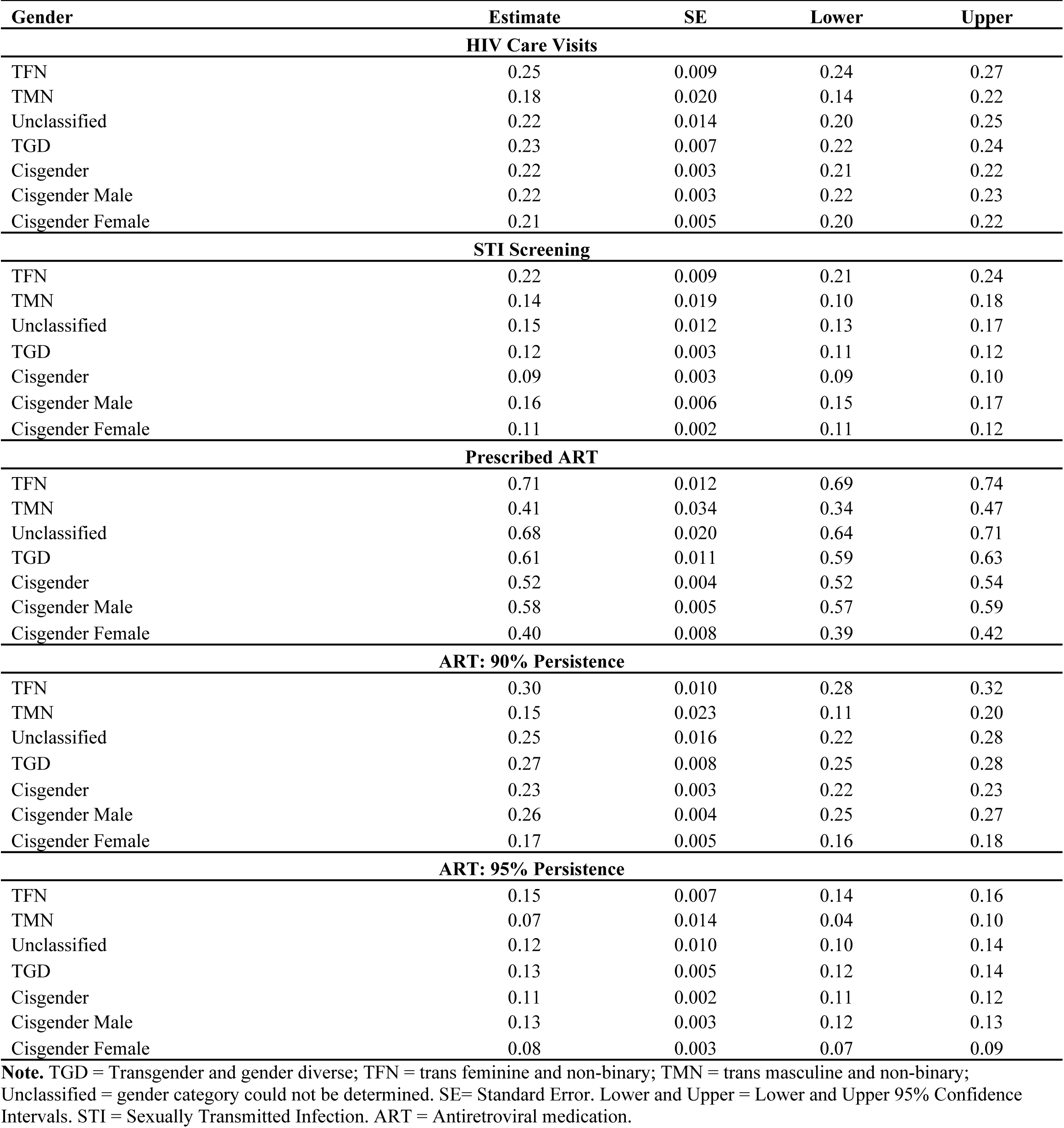
Predicted probabilities of HIV Care Continuum engagement among transgender and gender diverse (TGD) (n=1,678) and cisgender (n=10,681) Medicare beneficiaries with HIV, overall and by gender subgroup, 2008-2017.

| Gender | Estimate | SE | Lower | Upper |
| --- | --- | --- | --- | --- |
| **HIV Care Visits** |  |  |  |  |
| TFN | 0.25 | 0.009 | 0.24 | 0.27 |
| TMN | 0.18 | 0.020 | 0.14 | 0.22 |
| Unclassified | 0.22 | 0.014 | 0.20 | 0.25 |
| TGD | 0.23 | 0.007 | 0.22 | 0.24 |
| Cisgender | 0.22 | 0.003 | 0.21 | 0.22 |
| Cisgender Male | 0.22 | 0.003 | 0.22 | 0.23 |
| Cisgender Female | 0.21 | 0.005 | 0.20 | 0.22 |
| **STI Screening** |  |  |  |  |
| TFN | 0.22 | 0.009 | 0.21 | 0.24 |
| TMN | 0.14 | 0.019 | 0.10 | 0.18 |
| Unclassified | 0.15 | 0.012 | 0.13 | 0.17 |
| TGD | 0.12 | 0.003 | 0.11 | 0.12 |
| Cisgender | 0.09 | 0.003 | 0.09 | 0.10 |
| Cisgender Male | 0.16 | 0.006 | 0.15 | 0.17 |
| Cisgender Female | 0.11 | 0.002 | 0.11 | 0.12 |
| **Prescribed ART** |  |  |  |  |
| TFN | 0.71 | 0.012 | 0.69 | 0.74 |
| TMN | 0.41 | 0.034 | 0.34 | 0.47 |
| Unclassified | 0.68 | 0.020 | 0.64 | 0.71 |
| TGD | 0.61 | 0.011 | 0.59 | 0.63 |
| Cisgender | 0.52 | 0.004 | 0.52 | 0.54 |
| Cisgender Male | 0.58 | 0.005 | 0.57 | 0.59 |
| Cisgender Female | 0.40 | 0.008 | 0.39 | 0.42 |
| **ART: 90% Persistence** |  |  |  |  |
| TFN | 0.30 | 0.010 | 0.28 | 0.32 |
| TMN | 0.15 | 0.023 | 0.11 | 0.20 |
| Unclassified | 0.25 | 0.016 | 0.22 | 0.28 |
| TGD | 0.27 | 0.008 | 0.25 | 0.28 |
| Cisgender | 0.23 | 0.003 | 0.22 | 0.23 |
| Cisgender Male | 0.26 | 0.004 | 0.25 | 0.27 |
| Cisgender Female | 0.17 | 0.005 | 0.16 | 0.18 |
| **ART: 95% Persistence** |  |  |  |  |
| TFN | 0.15 | 0.007 | 0.14 | 0.16 |
| TMN | 0.07 | 0.014 | 0.04 | 0.10 |
| Unclassified | 0.12 | 0.010 | 0.10 | 0.14 |
| TGD | 0.13 | 0.005 | 0.12 | 0.14 |
| Cisgender | 0.11 | 0.002 | 0.11 | 0.12 |
| Cisgender Male | 0.13 | 0.003 | 0.12 | 0.13 |
| Cisgender Female | 0.08 | 0.003 | 0.07 | 0.09 |

**Note.** TGD = Transgender and gender diverse; TFN = trans feminine and non-binary; TMN = trans masculine and non-binary; Unclassified = gender category could not be determined. SE= Standard Error. Lower and Upper = Lower and Upper 95% Confidence Intervals. STI = Sexually Transmitted Infection. ART = Antiretroviral medication.

